# Epidemic indicators do not determine intervention performance

**DOI:** 10.64898/2026.03.27.26349564

**Authors:** Kris V Parag

## Abstract

Epidemic growth rates, reproduction numbers and counts of new infections are universally used to guide public health intervention decisions. It is widely and reasonably believed that larger values of these indicators evidence the need for more urgent or stringent control. Here we show that this intuition can fail dramatically. We construct pairs of epidemics with indistinguishable growth rates, reproduction numbers and infection curves but fundamentally divergent responses to identical interventions, with one epidemic subsiding while the other grows exponentially. Conversely, we identify pairs in which one epidemic exhibits larger indicators and causes three times as many infections, yet both become suppressed with equal effectiveness under the same intervention. These paradoxical outcomes arise from structural uncertainties in transmission, which are invisible to standard outbreak indicators but become decisive under feedback control. Because structural uncertainty is unavoidable when representing real outbreaks, epidemic controllability and intervention performance cannot be reliably inferred without explicitly modelling this feedback between transmission and intervention.

## 1. Introduction

Few would dispute that an epidemic with larger growth rate, higher reproduction number and greater count of infections warrants faster or stronger control action. Relatedly, epidemics that are indistinguishable from these indicators are universally expected to be equally controllable [1]. These reasonable premises underpin a vast body of epidemic modelling work that has helped to guide prospective public health intervention decisions or evaluate such decisions retrospectively [2, 3].

Most of this work models interventions (e.g., quarantines, lockdowns and vaccinations) as direct reductions to the reproduction number or growth rate [2, 4]. Infection trajectories (or associated case or death curves) are simulated under these assumed transmission reductions to compare or justify intervention strategies [3, 5]. Epidemic indicators, estimated prior to intervention, therefore serve as the basis for specifying and evaluating counterfactual transmission reductions.

We focus on two key features of this prevailing approach. First, it is open loop (OL) because testing counterfactuals under preset conditions typically disregards the feedback loop between control and transmission dynamics [6, 7]. Second, structural uncertainty arising from differences in model formulation, complexity and parametrisation mean that multiple transmission mechanisms can underlie the same epidemic indicators, blurring the association between projected infections and intervention effectiveness [8, 9].

Studies have examined these features separately, proposing closed-loop (CL) counterfactual designs (e.g., using model-predictive control) [6, 7] or identifying crucial uncertainties to resolve (e.g., via model averaging) [8, 9]. However, behavioural coupling, environmental drivers and other unquantifiable variables mean that interventions are, realistically and invariably, subject to structural uncertainty and feedback effects simultaneously. Here we expose how these interacting features can produce paradoxical and dramatic outcomes.

We find that feedback can amplify or attenuate structural uncertainty, rendering popular epidemic indicators that comprehensively describe OL growth, uninformative about CL dynamics [10]. Two counterintuitive phenomena emerge. First, epidemics indistinguishable in growth rate, reproduction number and infection trajectory can respond in fundamentally different ways to the same intervention. Second, an epidemic deemed more severe by every standard indicator may be no more difficult to control than one that appears milder.

## 2. Results

We model epidemics as renewal processes [1] involving cohorts of infectious individuals (see Methods). A cohort infected *s* days ago has infectiousness *Rw*(*s*), where *R* is the epidemic reproduction number and *w*(*s*) the probability of transmitting. The epidemic growth rate *r* depends on *R* and the generation time distribution, which is *w*(*s*) over all *s*. New infections on day *t, i*(*t*), grow if *R* > 1 and *r* > 1 (both correspond) [11]. Popular OL indicators for policy are *R, r* and the curve of *i*(*t*).

We simulate outbreaks with COVID-19 and Ebola virus parameters [3, 12]. Interventions remove age-dependent proportions *e*(*s*) of secondary infections generated by cohorts. This disrupts the transmission feedback loop, causing CL cohort infectiousness to converge to *Rw*(*s*)(1 − *e*(*s*)) (see Methods). As it is difficult to estimate *w*(*s*), substantial uncertainty often surrounds its structure and shape [13, 14]. We show that uncertainty in *w*(*s*) interacting with *e*(*s*) generically produces paradoxical outcomes under realistic intervention designs.

### 2.1. Identical epidemics, divergent responses

We construct approximate models (red) that preserve the mean generation time, *R* and *r* of the disease models (blue), while perturbing the shape of *w*(*s*) to represent structural uncertainty. In Fig 1 we simulate from these models, obtaining indistinguishable *i*(*t*) curves. From all OL indicators these are identical outbreaks that are equally controllable. However, we apply a simple control (similar to test-trace-isolate schemes [14]) and observe dramatic divergence. The approximate model is CL stable (*R* < 1, *r* < 1, decaying *i*(*t*)), but the original is uncontrolled and growing exponentially.

**Fig 1.**
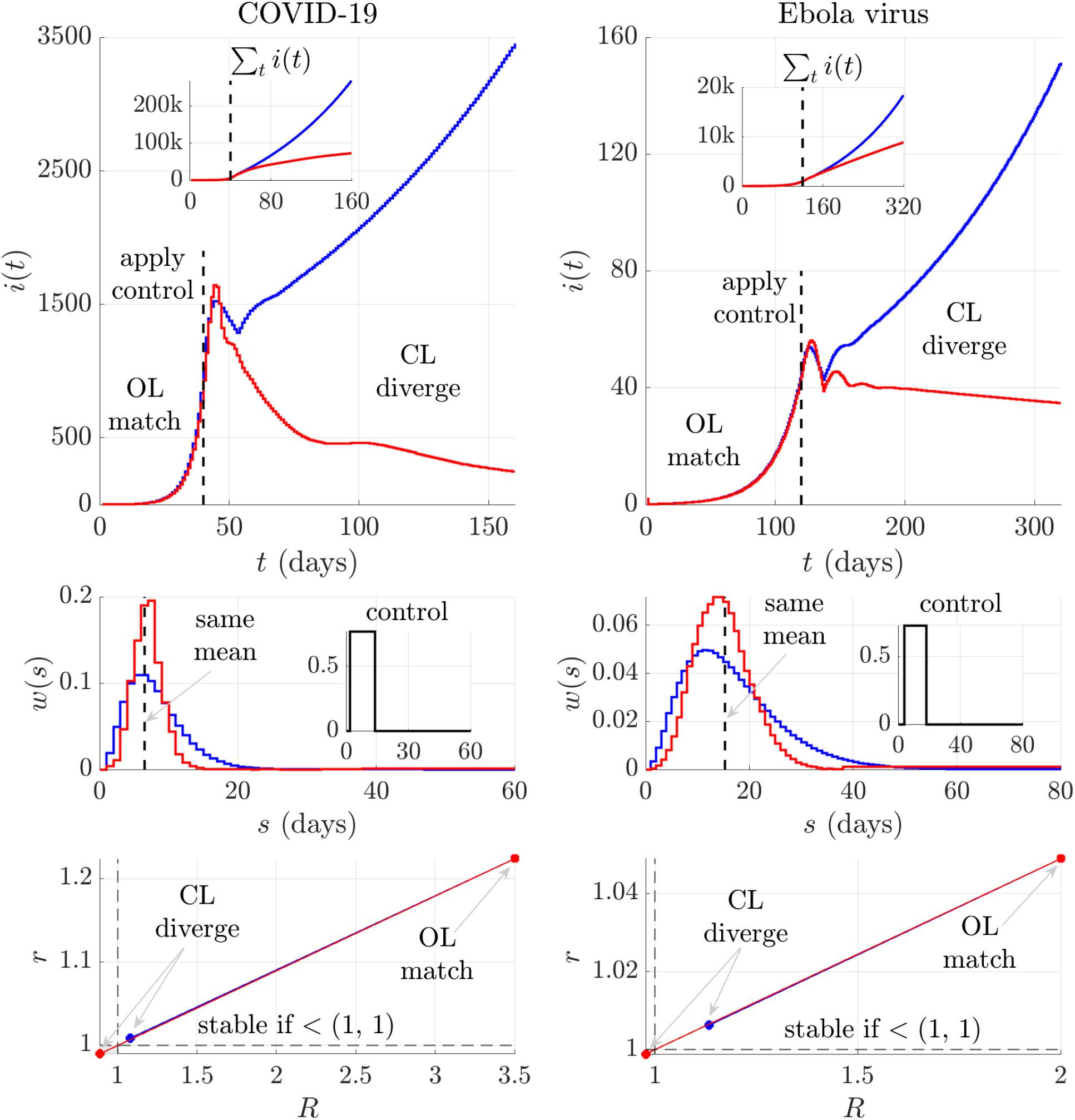
Indistinguishable epidemics but vastly divergent responses. We demonstrate that the same intervention applied to effectively identical epidemics can cause dramatically different transmission outcomes. We simulate (see Methods) under COVID-19 and Ebola virus generation time distributions (blue) and an approximate model for each disease (red). This model has exactly the same mean generation time, growth rate *r* and reproduction number *R* (bottom panels, squares) but different generation time distribution *w*(*s*) (middle panels). New infections *i*(*t*) are nearly identical (top panels, insets show cumulative infections) pre-intervention. By all conventional OL indicators, both epidemics are deemed equally controllable. We apply the same control (parametrised as proportions of infections removed, inset of middle panels) from the dashed timepoint. Surprisingly, it stabilises the approximate model but leaves the original disease model unstable. This is reflected in the resulting *r* and *R* (circles in bottom panels, shading is stable region) and the strikingly divergent CL trajectories of *i*(*t*).

### 2.2. Divergent epidemics, identical controllability

We formulate alternate models (red) with larger *R* and *r* than our disease models (blue) but similar *w*(*s*). In Fig 2 we see that these models have steeper growing *i*(*t*) with peaks ≈ 3 times that of the disease models. By every OL indicator, these alternate epidemics require more urgent or stringent control. However, we discover that both are equally controllable. A simple control action forces both models to indistinguishable *R* and *r* values. Small structural differences in the *w*(*s*), which caused the diverging *i*(*t*) become neutralised. Post-intervention both epidemic trajectories subside with matching rates.

**Fig 2.**
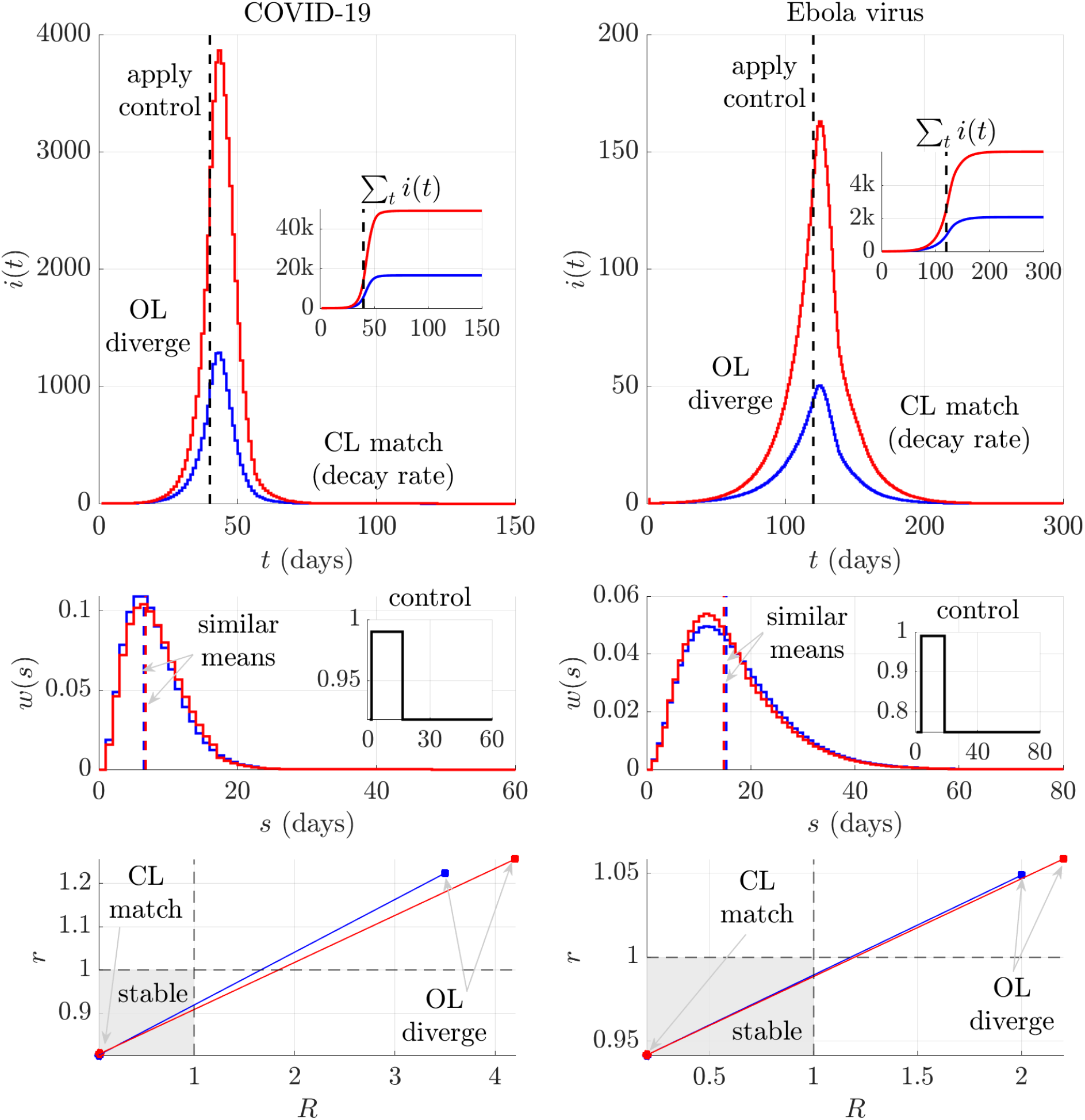
Vastly divergent epidemics with identical controllability. We demonstrate how the same intervention applied to remarkably different epidemics can yield indistinguishable transmission outcomes. We simulate (see Methods) under both COVID-19 and Ebola virus generation time distributions (blue) and an alternate model for each disease (red). This model has similar generation time distribution *w*(*s*) (middle panels) but a larger growth rate *r* and reproduction number *R* (bottom panels, squares). New infections *i*(*t*) (top panels with insets of cumulative infections) are tripled under the approximate model prior to intervening. By all conventional OL indicators, this model is substantially harder to control. We initiate the same control action (parametrised as proportions of infections removed, inset of middle panels) from the dashed timepoint. Surprisingly, it is equally effective, forcing both models in CL to matching *r, R* (circles in bottom panels, shading is stable region) and reducing *i*(*t*) at the same rate (with near identical log-differences in infections post intervention, not shown).

## 3. Discussion

The distinction between OL and CL responses is foundational to control theory [10] but underappreciated in epidemiology. Treating OL indicators and counterfactual shifts in those indicators as proxies for CL intervention performance is ubiquitous in epidemic modelling and elides this distinction. This conflation can be qualitatively misleading when control actions interact with structural uncertainty in transmission.

An identical intervention applied to outbreaks with indistinguishable OL growth indicators and infection trajectories can be unpredictable in effectiveness, suppressing some epidemics but failing to curb others (Fig 1). Conversely, the same intervention can be equally effective at controlling a slow, mild epidemic and a faster-growing one appearing more severe by every common indicator (Fig 2).

These paradoxical phenomena arise because feedback between control and transmission can magnify (Fig 1) or minimise (Fig 2) structural mismatches. This mechanism helps explain why interventions validated in models can underperform in real populations [1, 9]. Our findings suggest that widely-used epidemic indicators can mischaracterise intervention performance and hence obscure meaningful policy comparisons across regions, countries or socio-demographic groups, where structural heterogeneity is unavoidable.

We assumed homogeneous mixing, deterministic dynamics and static interventions to isolate the control-transmission feedback loop and identify fundamental OL-CL discrepancies. However, interventions also drive adaptations in population behaviour, spatial mobility and pathogen biology. These additional loops, together with associated heterogeneity and stochasticity, could exacerbate these discrepancies.

Given this complexity, resolving structural uncertainty via increasingly detailed OL modelling may be impractical [8]. A promising but under-explored alternative is to design interventions that remain provably robust to plausible structural perturbations. Control theory provides established tools for enforcing such robustness [10], although adapting them to realistic epidemic constraints is an open challenge. At minimum, wider recognition that current OL indicators are necessary but insufficient descriptors of CL outcomes is a first step to improving epidemic control and preparedness.

## 4. Materials and Methods

The renewal process [1] is widely used to model how new infections at time *t, i*(*t*), depend on past infections (over *s* < *t*) and infections *m*(*t*) seeded from external locations. This dependence is set by the reproduction number *R* and the generation time distribution, parametrised by *w*(*s*) as in Eq (1).

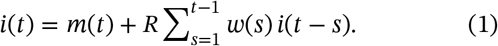

Here *w*(*s*) is the probability that an infection is transmitted in *s* time units. An individual generates *R* secondary infections while infectious. We assume homogeneous mixing and that the population size is large compared to total infections [1].

We reformulate Eq (1) as a structured population model (SPM), building on [11]. We structure by cohorts of infections so that **x**(*t*) = [*i*(*t*), *i*(*t* − 1), …]^′^ is our column-vector state. A cohort with age *s* has infectiousness *Rw*(*s*), which is its contribution to onward transmission. We capture this in **A** = [*Rw*(1), *Rw*(2), …] and define **B** = [1, 0, …]^′^, **T** as a lower shift matrix (a zero matrix with a sub-diagonal of 1s) and **m**(*t*) = [*m*(*t*), 0, …]^′^ to derive the SPM in Eq (2).

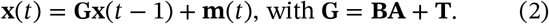

The first row of Eq (2) contains Eq (1). The eigenvalues *λ*[**G**] completely define the OL dynamics of the epidemic. The dominant eigenvalue is the discrete-time growth rate *r* = *λ*_max_[**G**]. We can also recover *R* as *λ*_max_[**G**(**I**−**T**)^−1^] with **I** as the identity matrix [11]. The signs of *r* − 1 and *R* − 1 correspond and if positive (negative) indicate a growing (waning) epidemic.

Eq (2) describes the positive feedback transmission loop linking past and new infections [15]. We model interventions as age-dependent disruptions to this loop. We remove a fraction 0 ≤ *e*(*s*) ≤ 1 of new infections caused by infected cohorts of age *s*. This captures a wide range of interventions. For example, a lockdown that halves spread has *e*(*s*) = 0.5 for all *s*. A test-trace-isolate scheme has *e*(*s*) > 0 for *a* ≤ *s* ≤ *b*, with *b* − *a* as an effective detection and isolation window [14].

When control is initiated, transient dynamics emerge as removals of infections by *e*(*s*) reshape the onward transmission of cohorts. After this transition, the epidemic again obeys a renewal process, but with reduced infectiousness *Rw*(*s*)(1 − *e*(*s*)) [14, 15]. This yields a modified Eq (2), in which state feedback reduces **G** by Δ = **B**[*Rw*(1)*e*(1), *Rw*(2)*e*(2), …]. The CL growth and reproduction indicators then satisfy Eq (3).

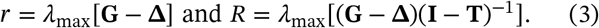

In the Results we examine the consequences of these shifted CL dynamics using parameters for COVID-19 and Ebola virus disease from [3, 12]. Matlab scripts reproducing all analyses are available at https://github.com/kpzoo/astrom.

We construct the approximate model in Fig 1 by solving an optimisation problem with hard constraints on preserving the *R, r* and mean generation time of the original disease model. This changes the *w*(*s*) shape only and captures structural uncertainty. We develop the alternative model in Fig 2 by redistributing some generation time mass to represent an uncertain but small second mode of transmission that only slightly perturbs the mean generation time. The settings and optimisation weights in our scripts can be varied to generate many instances of our paradoxical phenomena.

## Data Availability

No data were generated in this study. Matlab scripts reproducing all analyses are available at https://github.com/kpzoo/astrom.

## Notes

### Competing Interest Statement

The authors have declared no competing interest.

### Funding Statement

KVP acknowledges funding from the MRC Centre for Global Infectious Disease Analysis
(reference MR/X020258/1), funded by the UK Medical Research Council. This UK-funded grant is carried out in the frame of the Global Health EDCTP3 Joint Undertaking.

## References

1 C. Fraser et al., “Factors that make an infectious disease outbreak controllable”, Proc. Natl. Acad. Sci. U.S.A. 101, 6146–6151 (2004).

2 N. Haug et al., “Ranking the effectiveness of worldwide COVID-19 government interventions”, Nat. Hum. Behav. 4, 1303–1312 (2020).

3 S. Flaxman et al., “Estimating the effects of non-pharmaceutical interventions on COVID-19 in Europe”, Nature 584, 257–261 (2020).

4 C. M. Peak et al., “Comparing nonpharmaceutical interventions for containing emerging epidemics”, Proc. Natl. Acad. Sci. U.S.A. 114, 4023–4028 (2017).

5 R. Sonabend et al., “Non-pharmaceutical interventions, vaccination, and the SARS-CoV-2 delta variant in England: a mathematical modelling study”, Lancet 398, 1825–2835 (2021).

6 K. van Heusden et al., “Effective pandemic policy design through feedback does not need accurate predictions”, PLOS Glob. Public Health 3, e0000955 (2023).

7 S. Beregi et al., “EpiControl: a data-driven tool for optimising epidemic interventions and automating scenario planning to support real-time response”, medRxiv 2025.11.17.25340271 (2025).

8 S. Li et al., “Essential information: Uncertainty and optimal control of Ebola outbreaks”, Proc. Natl. Acad. Sci. U.S.A. 114, 5659–5664 (2017).

9 K. Shea et al., “Multiple models for outbreak decision support in the face of uncertainty”, Proc. Natl. Acad. Sci. U.S.A. 120, e2207537120 (2023).

10 K. J. Astrom and R. M. Murray, Feedback systems: an introduction for scientists and engineers, 2nd ed. (Princeton University Press, 2021).

11 B. Boldin, O. Diekmann, and J. A. J. Metz, “Population growth in discrete time: a renewal equation oriented survey”, J. Differ. Equ. Appl. 30, 1062–1090 (2024).

12 M. Van Kerkhove et al., “A review of epidemiological parameters from Ebola outbreaks to inform early public health decision-making”, Sci. Data 2, 150019 (2015).

13 K. V. Parag, B. J. Cowling, and B. C. Lambert, “Angular reproduction numbers improve estimates of transmissibility when disease generation times are misspecified or time-varying”, Proc. Roy. Soc. B 290, 20231664 (2023).

14 L. Ferretti et al., “Quantifying SARS-CoV-2 transmission suggests epidemic control with digital contact tracing”, Science 368, eabb6936 (2020).

15 K. V. Parag, “How to measure the controllability of an infectious disease?”, Phys. Rev. X. 14, 031041 (2024).

